## Supplementary figures and images for "From elimination to suppression: genomic epidemiology of a large Delta SARS-CoV-2 outbreak in Aotearoa New Zealand"

### Supplementary Figure 1

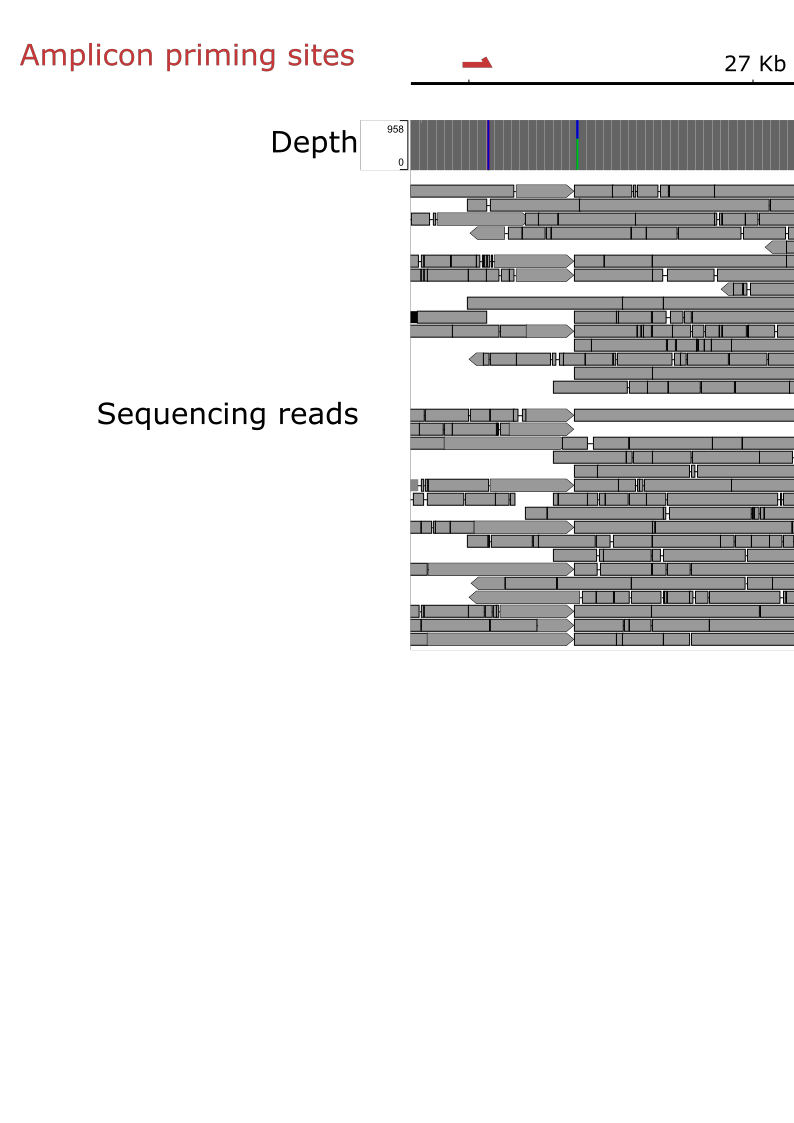
